# Autonomous Screening for Laser Photocoagulation in Fundus Images Using Deep Learning

**DOI:** 10.1101/2023.01.30.23285179

**Authors:** Idan Bressler, Rachelle Aviv, Danny Margalit, Yovel Rom, Sean Ianchulev, Zack Dvey-Aharon

**Affiliations:** AEYE Health, Inc.; New York Eye and Ear, Mount Sinai Hospital, NY

## Abstract

**Background:** Diabetic retinopathy is a leading cause of blindness in adults worldwide. AI with autonomous deep learning algorithms has been increasingly used in the analysis of retinal images particularly for the screening of referrable DR. An established treatment for proliferative DR is pan-retinal or focal laser photocoagulation. Training AI autonomous models to discern laser patterns can be important in disease management and follow-up.

**Methods:** A deep learning model was trained for laser treatment detection using the EyePACs dataset. Data was randomly assigned, by participant, into development (n= 18,945) and validation (n= 2,105) sets. Analysis was conducted at the single image, eye, and patient levels. The model was then used to filter input images for three independent AI models for various retinal indications, and changes in model efficacy were measured using AUC and MAE.

**Findings:** On the task of laser photocoagulation detection: AUC of 0.981 (CI 95% 0.971-0.87) was achieved at the patient level. AUC of 0.950 (CI 95% 0.943-0.956) was achieved at the image level. AUC of 0.979 (CI 95% 0.972-0.984) was achieved at the eye level.

When analyzing independent AI models, efficacy was shown to improve across the board on images of untreated eyes. DME detection on images with artifacts was AUC 0.932 (CI 95% 0.905-0.951) vs. AUC 0.955 (CI 95% 0.948-0.961) on those without. Participant sex detection on images with artifacts was AUC 0.872 (CI 95% 0.830-0.903) compared to AUC 0.922 (CI 95% 0.916-0.927) on those without. Participant age detection on images with artifacts was MAE 5.33 vs. MAE 3.81 on those without.

**Interpretation:** The proposed model for laser treatment detection achieved high performance on all analysis metrics and has been demonstrated to positively affect the efficacy of different AI models, suggesting that laser detection can generally improve AI powered applications for fundus images.

**Funding:** Provided by AEYE Health Inc.

## Introduction

Laser photocoagulation is a common and established procedure, in which laser pulses are used to coagulate retinal tissue, used to treat multiple retinal diseases [1]–[3]. Ablative photocoagulation is mostly used to prevent leakage and ischemic neovascularization in vascular retinal conditions such as diabetic retinopathy (DR) [4], [5], diabetic macular edema (DME) [6]– [8], retinal vein occlusion (RVO) [9], [10] and neovascular age-related macular degeneration (AMD) [11], [12].

Laser photocoagulation is generally divided into pan-retinal and focal; the former is delivered in the peripheral retina with deep ablative burns to stem the neovascular process [13], [14], while the latter is a lighter photocoagulative treatment delivered in the central macula to treat macular conditions [15], [16]. There are well established laser treatment protocols depending on disease severity and individual patient disease state [12], [17]–[19]. While laser photocoagulation is an effective treatment, it causes retinal scarring and is destructive to the retinal tissue leaving long term defects in the anatomy. [20]–[22].

Artificial Intelligence (AI) using fundus imaging has been increasingly employed in various ophthalmological applications [23], [24]. These applications include extraction of basic patient data, such as age and sex [25], detection of retinal pathologies, [26]–[28] and pathology development prediction [29]–[31]. AI methods rely on image pattern recognition, especially in areas in which the pathology is present. As such, laser photocoagulation may disrupt general pattern recognition by adding new patterns or artifacts, such as burns and scars, which the model is less trained to deal with. This is specifically problematic given that laser treatment is often done on areas of interest, such as leaky blood vessels, which are often the very areas that are most crucial to recognize.

The effect laser photocoagulation has on AI systems suggests that a tool to identify images of eyes which have undergone photocoagulation may be beneficial for the autonomous retinal-based diagnosis and follow-up treatment of patients. While previous methods of laser photocoagulation detection exist[32]–[36], this work, to the best of our knowledge, presents the first laser treated image detection method based on a large, diverse, and widely accepted database - in this case, EyePACS (https://www.eyepacs.org); the database contains images from a variety of manufacturers and patient populations, of varying image qualities.

## Methods

### Data

The data consisted of a subsample of the EyePACs dataset, which contains 45° angle fundus photography images and expert readings of said images. All images and data were de-identified according to the Health Insurance Portability and Accountability Act “Safe Harbor” before they were transferred to the researchers. Institutional Review Board exemption was obtained.

The dataset contained up to 6 images per patient visit: one macula centered image, one disc centered image, and one centered image, per eye. Each eye underwent expert reading, including but not limited to panretinal laser treatment presence, focal laser treatment presence, and image quality. All images of the subsample deemed readable by expert annotations were used.

The resulting dataset consisted of 21,050 images from 9,212 patients, of which 9,484 images (45%) had artifacts of panretinal laser treatment, 1,888 (9%) had artifacts of focal laser treatment, and 847 (4%) had both. This work combined focal and panretinal laser treatments into one category of laser treatment, resulting in an overall 10,525 (50%) images with laser treatment artifacts (**table 1**). Of these, roughly 77% of patients required dilation, where 54% of all patients received 1 gtt. tropicamide 1%, 17% received 1 gtt. tropicamide 0.5%, and 5% received other dilation agents.

**Table 1.**
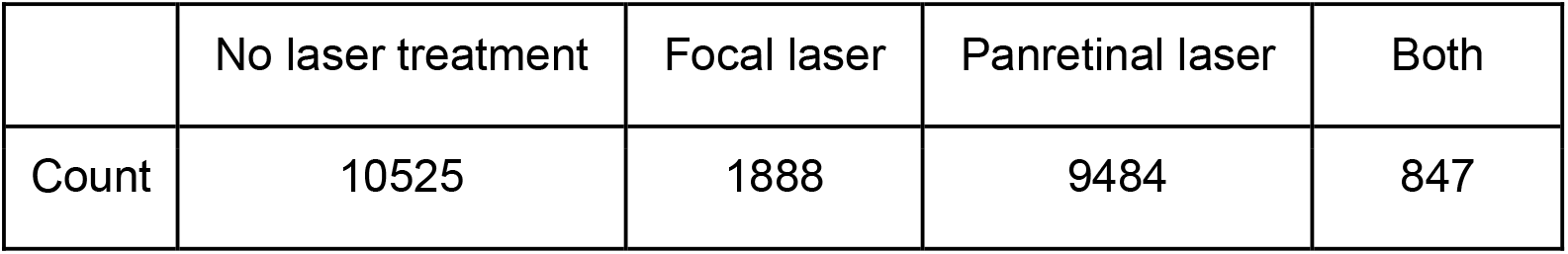
Laser treatment prevalence in the EyePACs dataset

|  | No laser treatment | Focal laser | Panretinal laser | Both |
| --- | --- | --- | --- | --- |
| Count | 10525 | 1888 | 9484 | 847 |

The average age of patients with laser treatment artifacts was 59.5 (10.0 SD) and 55% were female, compared to the patients who had not undergone laser treatment, for which the average age was 55.6 (11.3 SD) and 61% of which were female (**table 2**). The distribution of laser treatment images across DR levels is given in **table 3**; all laser treatment images were from patients with more than mild DR, and the majority were from patients with grade 4 DR.

**Table 2.** Patient demographics for patients who did and did not have laser treatment artifacts

|  | Count | Age (S.D) | Gender (% female) | Ethnicity (fraction) |
| --- | --- | --- | --- | --- |
| With laser | 10525 | 59.5 (10.0) | 51 | White = 0.59 (Hispanic = 0.95<br>non-Hispanic = 0.05)<br>Indian subcontinent origin = 0.11<br>African Descent = 0.07<br>Asian = 0.02<br>Not specified = 0.13<br>Other = 0.08 |
| No laser | 10525 | 54.7 (10.8) | 52 | White = 0.51 (Hispanic = 0.90<br>non-Hispanic = 0.10)<br>African Descent = 0.13<br>Indian subcontinent origin = 0.12<br>Asian = 0.03<br>Not specified = 0.14<br>Other = 0.07 |

**Table 3.** Distribution of laser treatment prevalence across different diabetic retinopathy grades

| DR grade | 0 | 1 | 2 | 3 | 4 | ungradable |
| --- | --- | --- | --- | --- | --- | --- |
| Total count | 558,451 | 52,928 | 106,377 | 16,909 | 19,470 | 25,718 |
| With laser | 0 | 0 | 365 | 175 | 9,607 | 136 |
| No laser | 558,451 | 52,928 | 105,742 | 16,734 | 9,863 | 25,582 |

### Quality assessment

A tool for image quality assessment was developed based on detecting visibility of fundus-specific characteristics. The given quality score for an image is an aggregation of the visibility from multiple areas within the fundus image. **Figure 2** demonstrates a few examples of images and their respective scores, showing the correlation between score and visual image quality.

This was done in order to remove low-quality images from the dataset, as the quality scores assigned by EyePACs are assigned to patients and not to individual images.

### Pre-Processing

Image pre-processing was performed in two steps for both datasets. Firstly, image backgrounds were cut along the convex hull, which contains the circular border between the image and the background. **Figure 1** shows an example of this process. Secondly, images were resized to 512×512 pixels. Lastly, using the aforementioned quality assessment tool, bad quality images were filtered out before training. The image quality threshold was set at the point at which model performances were not improved by filtering additional images, resulting in 1,373 images filtered, approximately 6.5% of the data.

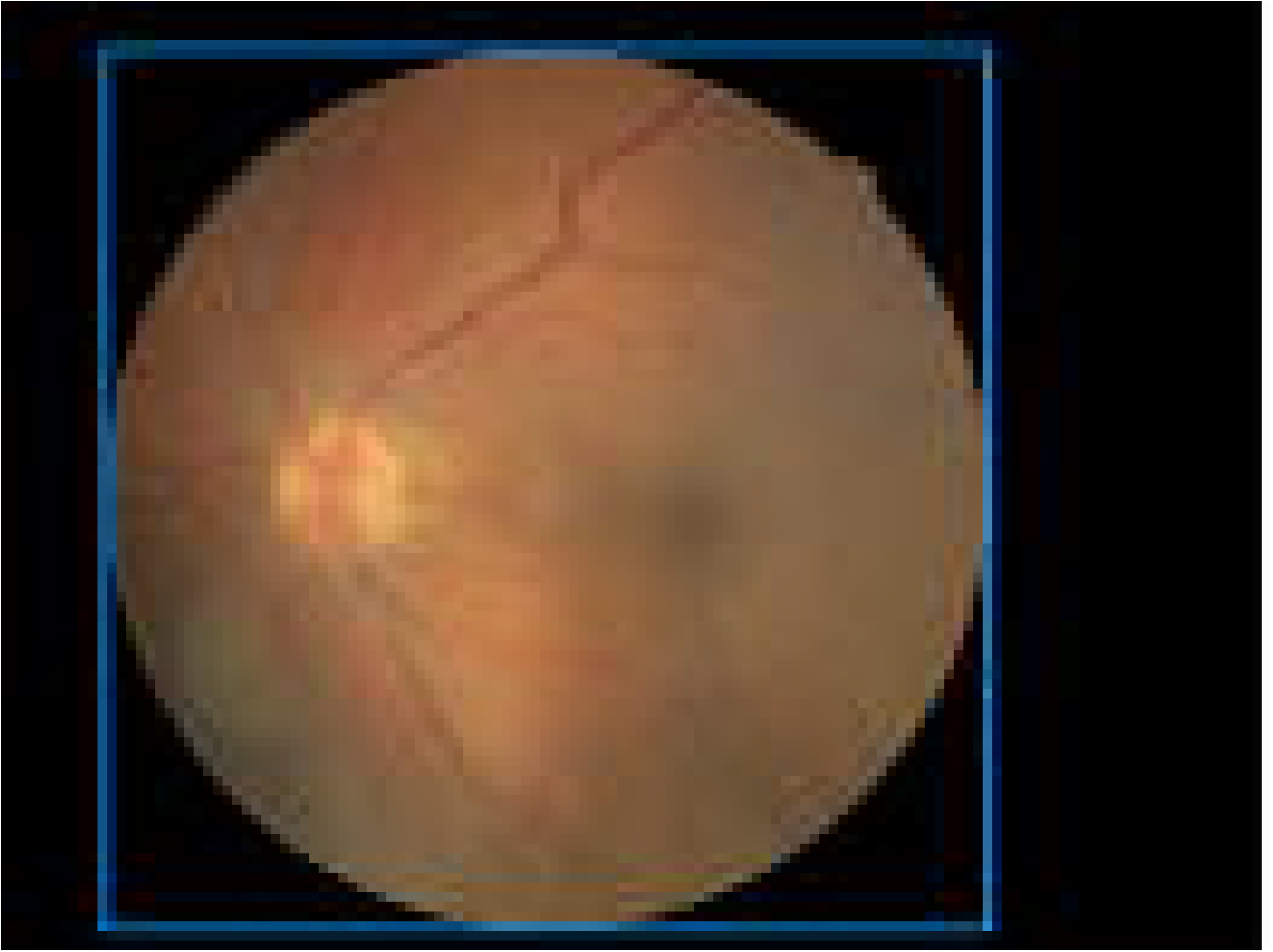

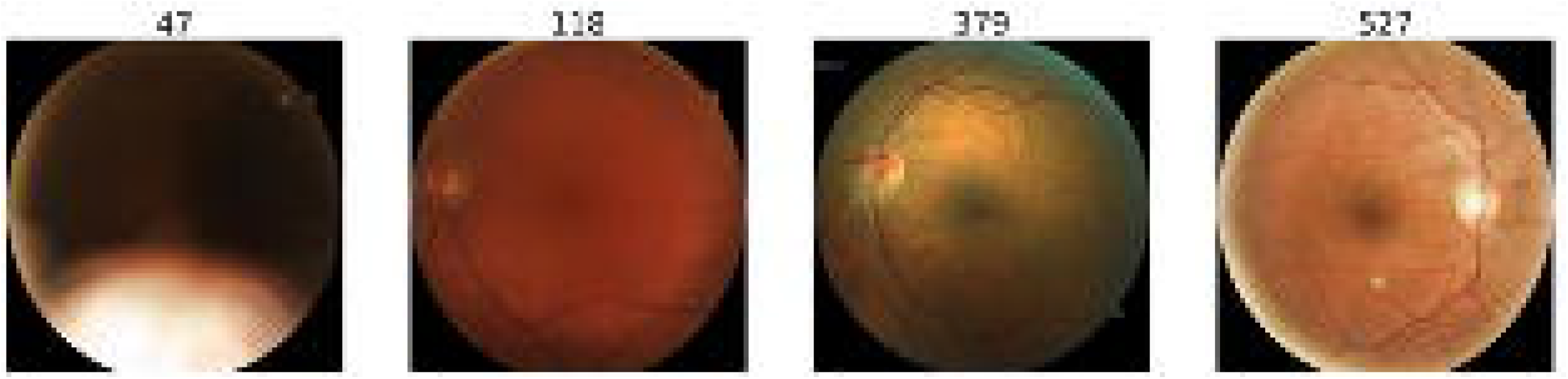

### Model training

The data was then divided into training, validation, and test datasets at a ratio of 80%, 10%, and 10% respectively. A binary classification neural network was trained. The model architecture was automatically fitted to best balance the model performance vs. model complexity tradeoff.

Hyperparameter tuning was done on the validation set.

### Statistical analysis

The metrics used for model assessment were accuracy, sensitivity, specificity, and area under the receiver operating characteristic curve (AUC). For each metric the bias corrected and accelerated bootstrap method [37] was used to produce a 95% confidence interval.

### Analysis levels

Laser detection was done on three different levels. The first, detection on the individual image level, was the basic task for which the model was trained. The second, detection on the eye level, used all images from a given eye and the image for which the model had the highest degree of confidence was selected for analysis. For the third, detection on the patient level, the results from both eyes were compared and the eye with the higher model confidence was selected to produce a patient-level result.

### Effect on imaging tasks

The effect that laser treatment has on imaging tasks was measured by applying the laser detection model as a preprocessing step for a model for the detection of DME, which was developed based on the EyePACs dataset [38], and a model for age detection, also developed based on the EyePACs dataset.

The performance on these tasks was measured in AUC on a separate validation set containing images both with and without laser treatment artifacts. The 95% confidence interval was calculated using the accelerated bootstrap method for each population and compared for significance.

A regression model was additionally trained for age detection, and the mean absolute error (MAE) between the patient’s age and predicted age was calculated on a separate validation set. The validation set was separated into patients with and without laser treatment artifacts, such that the mean age between these populations was the same. Significance in mean absolute error between the two populations was calculated using a student’s T-test. Detailed patient statistics of these experiments are given in **supplementary table 1**.

## Results

The results for the different analysis methods of laser artifact detection were as follows (**table 4**): on the image level, sensitivity of 0.883 (CI 95% 0.868, 0.897) and specificity of 0.880 (CI 95% 0.864, 0.894) were achieved. On the eye level, sensitivity of 0.925 (CI 95% 0.900, 0.945) and specificity of 0.931 (CI 95% 0.916, 0.944) were achieved. On the patient level, sensitivity of 0.929 (CI 95% 0.881, 0.947) and specificity of 0.926 (CI 95% 0.911, 0.944) were achieved.

**Table 4.**
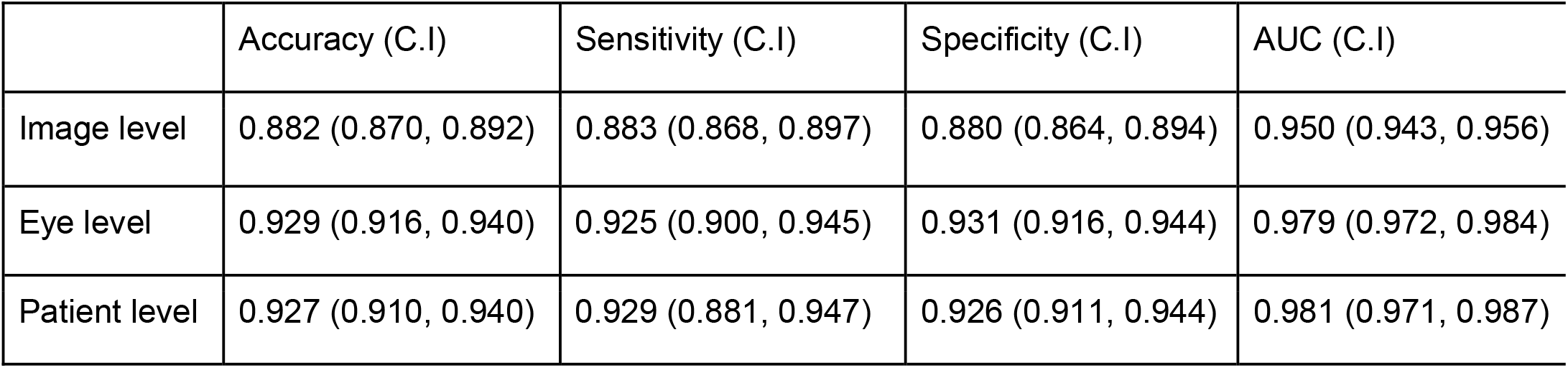
Laser treatment detection results on the EyePACs dataset for the three analysis levels performed, given in accuracy, sensitivity, specificity, and AUC with a 95% confidence interval. CI noted in parentheses.

|  | Accuracy (C.I) | Sensitivity (C.I) | Specificity (C.I) | AUC (C.I) |
| --- | --- | --- | --- | --- |
| Image level | 0.882 (0.870, 0.892) | 0.883 (0.868, 0.897) | 0.880 (0.864, 0.894) | 0.950 (0.943, 0.956) |
| Eye level | 0.929 (0.916, 0.940) | 0.925 (0.900, 0.945) | 0.931 (0.916, 0.944) | 0.979 (0.972, 0.984) |
| Patient level | 0.927 (0.910, 0.940) | 0.929 (0.881, 0.947) | 0.926 (0.911, 0.944) | 0.981 (0.971, 0.987) |

The results of laser artifact detection for each DR level are displayed in **table 5**: the model achieved 0.910 AUC (CI 95% 0.866, 0.941) for DR level 2, 0.887 AUC (CI 95% 0.758, 0.954) for DR level 3, 0.929 AUC (CI 95% 0.918, 0.938) for DR level 4 and 0.772 AUC (CI 95% 0.904, 0.968) for ungradable DR level. DR levels 0 and 1 did not have any laser treated examples, thus most metrics are not defined for these groups.

**Table 5.** Results on the EyePACs dataset across DR grades, given in accuracy, sensitivity, specificity, and AUC with a 95% confidence interval. CI noted in parentheses.

| DR grade | 2 | 3 | 4 | Ungradable |
| --- | --- | --- | --- | --- |
| Sensitivity (C.I) | 0.680 (0.591, 0.756) | 0.667 (0.472, 0.806) | 0.869 (0.853, 0.884) | 0.846 (0.652, 0.957) |
| Specificity (C.I) | 0.958 (0.906, 0.984) | 0.960 (0.791, 1) | 0.845 (0.821, 0.866) | 0.688 (0.400, 0.882) |
| AUC (C.I) | 0.910 (0.866, 0.941) | 0.887 (0.758, 0.954) | 0.929 (0.918, 0.938) | 0.772 (0.904, 0.968) |

**Table 6** shows the difference in results in laser artifact detection between patients with and without DME. The model achieved 0.955 AUC (0.948, 0.962) for non DME patients vs. 0.908 AUC (0.884, 0.927) for DME patients, demonstrating that these conditions do affect results, but the model achieves high performance irrespective of them.

**Table 6.**
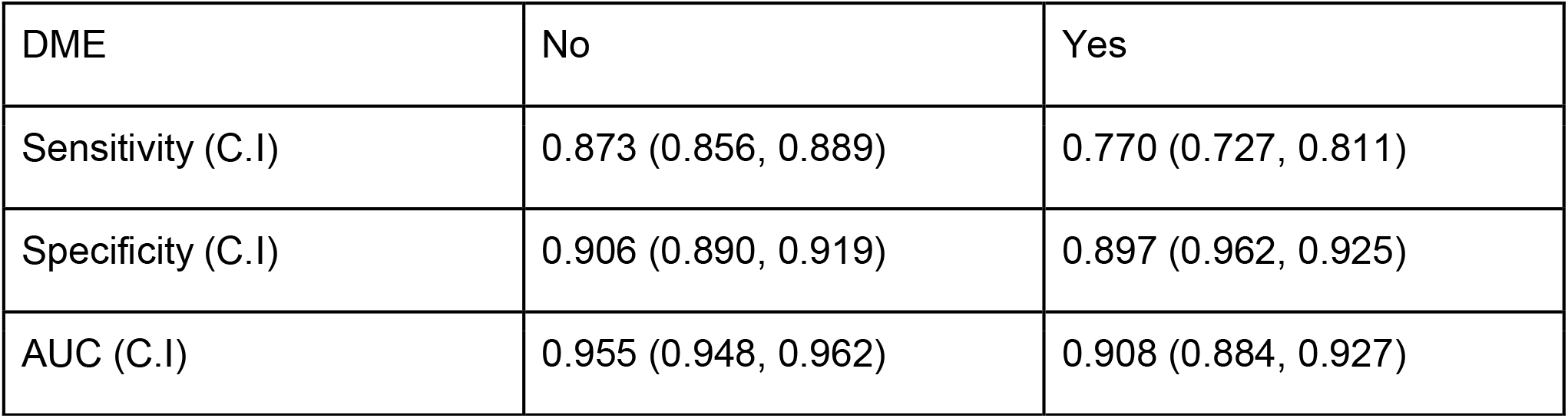
Results on the EyePACs dataset for patients with and without DME, given in accuracy, sensitivity, specificity, and AUC with a 95% confidence interval. CI noted in parentheses.

| DME | No | Yes |
| --- | --- | --- |
| Sensitivity (C.I) | 0.873 (0.856, 0.889) | 0.770 (0.727, 0.811) |
| Specificity (C.I) | 0.906 (0.890, 0.919) | 0.897 (0.962, 0.925) |
| AUC (C.I) | 0.955 (0.948, 0.962) | 0.908 (0.884, 0.927) |

**Table 7** displays the results of laser artifact detection for images which passed (high quality) and didn’t pass (low quality) the quality filter, showing a significant difference between the populations. The results for low quality images, which were filtered out, were 0.787 sensitivity (CI 95% 0.710, 0.849), 0.793 specificity (CI 95% 0.709, 0.860), and 0.857 AUC (CI 95% 0.803, 0.898); compared to 0.854 sensitivity (CI 95% 0.838, 0.869), 0.904 specificity (CI 95% 0.890, 0.917), and 0.948 AUC (CI 95% 0.941, 0.955) for high quality images which passed the filter.

**Table 7.** Results for images who were filtered out and not filtered out by the image quality tool, given in accuracy, sensitivity, specificity and AUC with a 95% confidence interval

|  | Sensitivity (C.I) | Specificity (C.I) | AUC (C.I) |
| --- | --- | --- | --- |
| Filtered out | 0.787 (0.710, 0.849) | 0.793 (0.709, 0.860) | 0.857 (0.803, 0.898) |
| Remained | 0.854 (0.838, 0.869) | 0.904 (0.890, 0.917) | 0.948 (0.941, 0.955) |

The effect of laser detection and subsequent filtration on the aforementioned three tasks of DME detection, age prediction, and sex detection, were as follows: DME detection results for images with no laser artifacts were 0.955 AUC (CI 95% 0.948, 0.961), compared to images with laser artifacts, on which the model achieved 0.932 AUC (CI 95% 0.905, 0.951). Age prediction results for images with no laser artifacts, after age adjustment, were 3.81 mean absolute error (MAE), compared to images with laser artifacts, on which the model achieved 5.33 MAE. T-test analysis shows a significance of p < 1e-4. Sex detection results for images with no laser artifacts were 0.922 AUC (CI 95% 0.916, 927), compared to images with laser artifacts for which the model achieved 0.872 AUC (CI 95% 0.830, 0.903).

The aggregation of these results is shown in **table 8**.

**Table 8.**
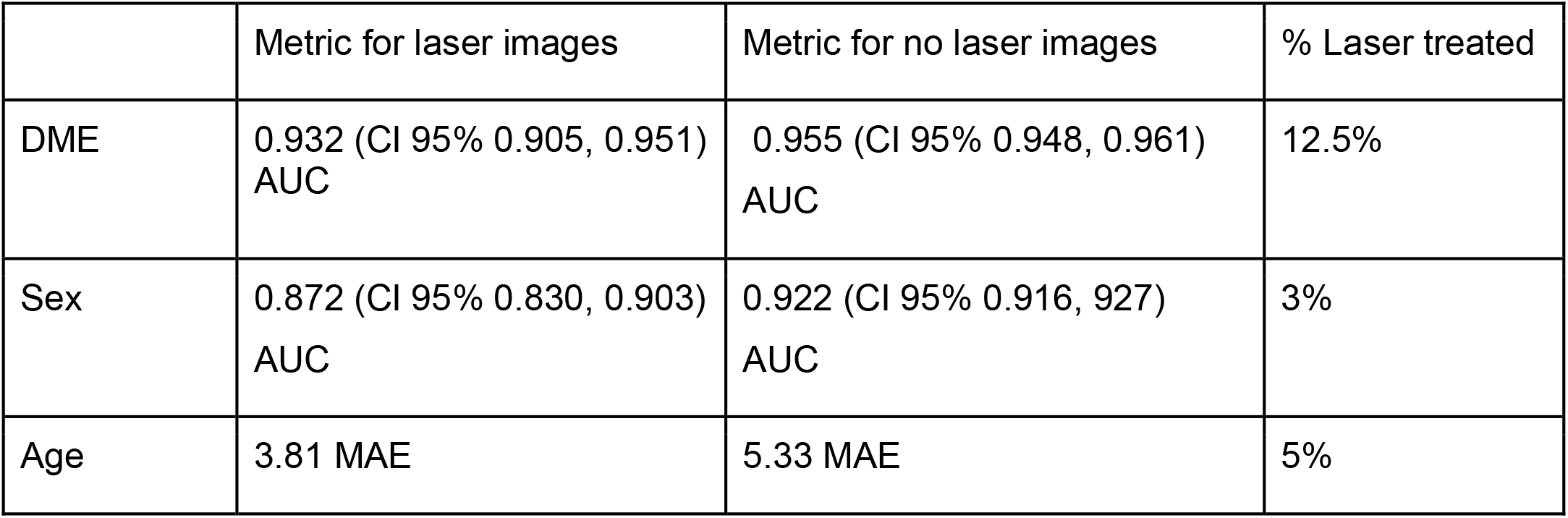
Results for the three experiments conducted for the effect of laser treatment, showing the results in terms of AUC for sex and DME detection and mean average error for age detection. The percentage of filtered images is shown.

|  | Metric for laser images | Metric for no laser images | % Laser treated |
| --- | --- | --- | --- |
| DME | 0.932 (CI 95% 0.905, 0.951)<br>AUC | 0.955 (CI 95% 0.948, 0.961)<br>AUC | 12.5% |
| Sex | 0.872 (CI 95% 0.830, 0.903)<br>AUC | 0.922 (CI 95% 0.916, 927)<br>AUC | 3% |
| Age | 3.81 MAE | 5.33 MAE | 5% |

## Discussion

This work proposed a method for the automatic detection of laser treatment artifacts in fundus images, which may also serve as a component in the future development of AI systems for different diagnoses based on retinal imaging. Such tasks may need to consider images of laser-treated eyes differently from non-treated eyes according to their design needs; some may choose to discard these images, while others may analyze them in a manner differently to images of untreated eyes. Accordingly, and in accordance with the degree to which laser treatment affects the task in question, the proposed system may be used at different operating points with different sensitivity-specificity balances. Discarding laser-treated images is a viable option for most automated retinal screening applications, as these patients should already have an awareness of the need for regular screening.

Previous studies on the autonomous detection of laser burns from fundus images have been on a smaller scale (roughly 2 orders of magnitude) [32]–[36]. The importance of scale is in the better representation of real-life conditions; specifically, this study allows better representation of various image qualities, camera manufacturers, and populations. Additionally, a wider range of clinical conditions, such as DR and DME, are represented in this study both with and without laser treatment, and the proposed system shows high performance across these conditions.

The effect laser treatment has on imaging tasks, and the model’s ability to detect relevant images, were validated by checking the model’s effect on different AI tasks involving retinal images. A significant difference was found for all three tasks, showing the relevance of the proposed method for future AI tasks.

A limitation of this work is the lack of differentiation between focal and panretinal laser treatments that were grouped as one in this work. Future works may differentiate between the two, given increased data. Furthermore, even though the base characteristics of laser photocoagulation remain similar across conditions, the addition of AMD-specific databases to the training set may improve results.

In addition, and in the same vein of the presented work, machine learning methods to detect patients with DME who will require future laser treatment may be developed. This would require training a model, similar to the one presented, on a dataset generated from a longitudinal study tracking the progression of patients with diabetes.

## Data Availability

All data produced in the present study are owned by AEYE Health, Inc.

