## supplementary table 1 for "Autonomous Screening for Laser Photocoagulation in Fundus Images Using Deep Learning"

| Task | Patients | Images | Mean Age (S.D) | Gender | Ethnicity (fraction) |
| --- | --- | --- | --- | --- | --- |
| DME detection | 1,607 | 3,576 | 54.80 (10.35) | 50% female | white = 0.55 (Hispanic = 0.93, non-Hispanic = 0.07)  ethnicity not specified = 0.13  Indian subcontinent origin = 0.12  African Descent = 0.11  Asian = 0.02  Other = 0.07 |
| Age | 2612 | 4754 | 61.78 (10.88) | 57% female | white = 0.50 (Hispanic = 0.88, non-Hispanic = 0.12)  ethnicity not specified = 0.17  African Descent = 0.13  Indian subcontinent origin = 0.08  Asian = 0.03  Other = 0.09 |
| Sex | 1318 | 10506 | 54.94 (11.06) | 59% female | white = 0.60 (Hispanic = 0.83, non-Hispanic = 0.17)  ethnicity not specified = 0.14  African Descent = 0.09  Indian subcontinent origin = 0.06  Asian = 0.03  Other = 0.08 |

**Supplementary**

Table 1. Cohort statistics for the three modules tested for laser treatment effect.
